# Telemedical communication patterns in myasthenia gravis in a remote monitoring study

**DOI:** 10.64898/2026.01.19.26344272

**Authors:** Meret Herdick, Sophie Lehnerer, Maximilian Mönch, Lea Gerischer, David Legg, Haoqi Sun, Pushpa Narayanaswami, Andreas Meisel, Maike Stein

## Abstract

**Purpose:** Myasthenia gravis (MG) is a rare neuromuscular disease. In-person appointments in specialized centers are not readily available, especially on short notice. The purpose of this study was to analyze patient-specialist communication through a telemedical platform.

**Methods:** In a randomized controlled study 45 MG patients were observed over three months. The intervention group (N=30) was monitored via a mobile application (‘app’) that enabled chat function and assessed MG-specific outcome measures and data from external devices. We quantitatively evaluated communication patterns for patients and specialists from the chat. Controls (N=15) received standard of care. Perceived MG specialist accessibility and contact with physicians outside the study team regarding MG-related issues was assessed in both groups.

**Findings:** In total, MG specialists were contacted a median of six times per patient via the chat concerning medical (54.5%), technical (25.3%), organizational (19.1%), and other (1.1%) topics. Specialists initiated contact a median of eight times per patient, most frequently addressing medical concerns (49.4%), 38.4% of which were recommendations for medication changes. Specialists also addressed technical (40.0%), organizational (9.4%) and other (1.3%) issues. Contacts with physicians outside the study team did not differ significantly between groups. Perceived specialist accessibility was rated significantly higher with telemedical support by the intervention group.

**Conclusions:** MG patients have a significant demand for timely advice especially concerning treatment recommendations. Future studies should evaluate the potential of telemedical solutions to prevent worsening of symptoms.

**Trial registration:** The study was registered under DRKS00029907 on August 19, 2022.

## 2 Introduction

Myasthenia gravis (MG) is a rare, autoimmune disorder characterized by an exercise dependent muscle weakness, which can affect all voluntary muscle and can potentially lead to myasthenic crises.^1^

In Germany, people with MG are often treated in specialized integrated Myasthenia Centers (iMZ), certified by the patient organization, German Myasthenia Association (DMG). MG patients may require frequent clinical visits to make treatment decisions, but availability of in-person consultations is limited by several factors, including scheduling and flexibility of iMZ consultation times, and patient-related challenges such as travel distance and/or transportation issues.^2–5^ In a study analyzing >1,700 email counselling requests from MG patients to iMZ Berlin and the DMG, more than half of inquiries pertained to medical issues highlighting a perceived need for timely advice, but requests also covered topics such as organizational or social legislation issues (e.g., rehabilitation, medical remedies and aids).^6^

In recent years there has been a surge in the development of application (‘app’) based digital solutions for patients, e.g. for headache disorders^7^ or Parkinson’s disease^8, 9^, to monitor symptoms. Digital health care solutions that enable monitoring in addition to a communication feature for patient-specialist interaction may be of particular value for patients with MG due to the fluctuating nature of the disease.

The aim of this analysis is to examine MG-patient-specialist communication via chat supported by remote monitoring data. In addition, perceived accessibility of MG specialist care and contacts with other physicians regarding MG-related issues is investigated.

## 3 Materials and Methods

### 3.1 Study design

The study was performed over a period of 12 weeks between 04/2023 and 09/2023. Detailed description of study protocol has been published previously.^10^

Patients aged 18 and above with a diagnosis of MG defined as a documented international statistical classification of diseases and related health problems 10th revision (ICD-10) code of G70.0 for at least six months prior to study enrolment treated in the iMZ Berlin were eligible for participation. Diagnosis of MG was based on clinical features of fatigable muscle weakness supported by at least one of the following: antibody positivity, electrophysiological testing (detection of decrement or increased jitter) and/or clear response to anticholinesterase inhibitors. Exclusion criteria included organic, including symptomatic, mental disorders (ICD-10: F00-F09), intellectual disability (ICD-10: F70-F79), and other somatic, psychiatric or neurological disorders or treatments that could potentially affect measured parameters and cause an inability to use a smartphone to enter data. Patients were recruited through the distribution of flyers and during in-person appointments at the iMZ Berlin.

Randomization was stratified based on sex and the Myasthenia gravis foundation of America (MGFA) status classification at the time of enrolment. The intervention group received telemedical services through the platform ‘MyaLink’ that comprises a patient app and a web-based platform for specialist use in addition to standard of care, whereas the control group received only standard of care. In this context, standard of care refers to routine outpatient visits scheduled well in advance; there was no standardized or structured way for patients in the control group to contact the center between visits, except by presenting to the hospital emergency department in case of clinical deterioration. Unlike the intervention group, control group patients did not receive any remote clinical reviews (i.e. telemedical check-ups) or structured remote monitoring during the study period.

### 3.2 Data collection

Standard of care included a baseline and end-of-study visit three months apart conducted at the study center consisting of an in-person appointment with a specialist from the study team and assessment of study relevant details including medical history, medication, hospitalizations, care-related questions (e.g. healthcare provider and their accessibility, areas requiring improvement in MG care, and perceived level of personal influence over MG), and various PROMs (Patient-Reported Outcome Measures). PROMs assessed were MG-ADL (Myasthenia gravis activities of daily living profile; 0-24 point scale),^11^ MG-QoL15r (Myasthenia gravis quality of life 15 item, revised; 0-30 point scale),^12^ SSQ (single simple question, 0-100%),^13^ PASS (patient-acceptable symptom state; “yes” or “no” response),^14, 15^ HADS (Hospital anxiety and depression scale; 0-42 point scale),^16–18^ and CFS (Chalder Fatigue Scale; 0-33 point scale).^13, 19, 20^ MG-specific clinical assessment was conducted using the Quantitative Myasthenia gravis Score (QMG; 0-39 point scale).^21^

In addition to standard of care, intervention group patients were able to contact specialists via the MyaLink chat function at any time throughout the study. The intervention group regularly assessed PROMs through the app and monitored vital parameters with external devices. These were an activity tracker (Garmin Vivosmart 5) which assessed heart rate, oxygen saturation, and step count and a digital spirometer (MIR Spirobank Smart One) for measurement of forced vital capacity. Data from both external devices were directly incorporated in the MyaLink app via bluetooth. Additionally, the single breath count test (SBCT) was performed by intervention group patients.^22^ At the baseline visit patients from the intervention group were instructed on the usage of MyaLink and external devices. In week four and week eight specialists performed a telemedical check-up (TCU) i.e. a systematic review of remote monitoring data and, if necessary, initiated contact via the chat (e.g., for medication changes). In between TCUs interactions occurred if patients reached out via the MyaLink chat. Specialists provided a response and where necessary, monitoring parameters were further reviewed based on the chat content.

Furthermore, we quantitatively assessed contact with other physicians not associated with our center regarding MG-related aspects in both groups. During baseline visits all study participants were questioned regarding the perceived availability of their MG specialist. At the end-of-study visit, patients in the intervention group were questioned again regarding the perceived availability of their MG specialist.

The study team consisted of five members, including three neuromuscular specialists (physicians with five to ten years of experience in MG care). Specialists performed in-person baseline and end of study visits in both groups and reviewed the chat in the intervention group aiming to answer within 24 hours on weekdays. A study nurse instructed intervention group patients in the use of MyaLink and external devices at the baseline visit. A research assistant reviewed data entry and was responsible for quality control.

### 3.3 Definitions and categorizations of patient-specialist communication patterns

Patients in the intervention group and specialists were able to contact each other through the MyaLink chat at any time. If the initial contact and subsequent responses pertained to the same topic, the interaction could extend over multiple days, but was still considered a single contact. However, if a new issue or question arose during an exchange, it was classified as a new contact. The content of contacts was categorized into pre-defined topics based on personal experience and previously published analyses on MG patient inquries. ^6^ In the event of ambiguities regarding categorization of an inquiry and its topic, cases were reviewed by another specialist from the study team to ensure interrater reliability.

The content of patient-initiated contacts was categorized into the following main topics: *medical issues* (all medical inquiries)*, social/legislation inquiries* (e.g. in regard to communication with authorities, rehabilitation, aids and remedies)*, organizational inquiries* (e.g. appointment scheduling issues)*, technical inquiries* (concerning the use of MyaLink and wearable devices) and *others*. If a contact involved more than one topic, multiple topics were selected accordingly. Some topics allowed for further subcategorization i.e. *medical issues* were divided into requests concerning *MG-related therapy, myasthenic complaints, side effects of medication* and *other medical reasons*. *Technical inquiries* were further divided into *initial setup questions* (questions within the first few days after study start e.g. for connecting external devices to MyaLink) and *technical questions*.

Content of specialist-initiated contacts was divided into topics addressed at TCUs and topics addressed in between TCUs. Reasons for contacts and recommendations from the specialist were recorded and categorized into different topics both at TCUs and in between TCUs. For TCUs, main topics were categorized into *medical issues, technical issues* and *others*. In between the TCUs main topics were categorized into *medical issues, technical issues,* and *organizational issues*. Both at TCUs and in between TCUs *medical issues* were further divided into subtopics with direct medical consequences including *change in medication* (i.e. recommendations for medication changes)*, referral to outpatient specialist/further diagnostics, referral to emergency department/hospital, laboratory check, increase in monitoring frequency* and *change in behavior (e.g. lifestyle changes)* as well as general medical advice: *Consultation about clinical complaints, deterioration of monitoring data, side effects of medication,* and *other medical reasons (non-MG related)* (only applicable in between TCUs). *Technical issues* were divided into the following subtopics: *initial setup, reminder for missing data entry* (i.e. if assigned PROMs or external device measurements were not performed), and *technical recommendations* (only applicable at TCUs).

At TCUs monitoring parameters that triggered specialist-initiated contact were recorded including *information based on chat interactions, medical reports and documents (uploaded in the app), PROMs* (MG-ADL, MG-QoL15r, SSQ, PASS, HADS, CFS), *wearables (heart rate, oxygen saturation, step count), spirometry (forced vital capacity)* and *SBCT*.

### 3.4 Statistical analysis

The primary endpoint of this study was feasibility (measured by adherence rates for digital PROMs assessment) and usability and has previously been published.^23^ Next to the here presented secondary endpoint analysis of communication patterns further research questions focusing on clinical endpoints will be presented in separate forthcoming publications:

1) Clinical endpoints: Does digital care with MyaLink reduce the disease severity of MG patients? Can MyaLink help to improve the quality of life of MG patients?
2) Prediction of MG worsening (‘digital biomarker’): Can telemonitoring and wearable data be valuable to predict impending crises and exacerbations in patients with MG?
3) Remote assessment of respiratory function: Is the single breath count test a feasible tool for remote assessment of respiratory function in MG?

Next to data from the MyaLink platform (exported as .csv) all data were stored in a REDCap ^®^ (Research Electronic Data Capture, version 13.7.31) database. The statistical analyses of the herein presented results were performed using R statistical software (version 4.2.2).^24^ Descriptive statistics (median, interquartile range, minimum, maximum), nonparametric tests, and effect sizes for continuous variables were selected after checking whether the data followed a Gaussian distribution graphically and double-checking with the Shapiro-Wilk test. Thus, the Mann-Whitney U test with Cliff’s Delta as the effect size was used for group comparisons of age and disease duration as well as MG-ADL and QMG scores.^25^ Categorical data are presented as absolute and relative frequencies, and group differences in proportions were assessed with Chi-squared tests. Depending on the number of categories, the strength of association between group membership and categorical outcome was assessed using odds ratios or Cramér’s V as effect sizes. Odds ratios were calculated for sex, MG therapies, and contacts with other physicians, and Cramér’s V was used for MGFA status and contact specifications. The analyses of the subjective availability of specialized medical services for the treatment of MG were split because only the intervention group had data at the end of the study. Therefore, responses were analyzed descriptively using bar graphs. Differences in response proportions between groups at baseline were examined using the Chi-squared test and Cramér’s V as the association effect size. Finally, the change in subjective availability for the intervention group from baseline to the end of the study was analyzed using Cochran’s Q test with η^2^ as the effect size measure for multiple paired binary variables.^26^ Due to the exploratory nature of this study no adjustments for multiple testing were necessary.

### 3.5 Standard protocol approvals, registrations, and patient consent

This study received approval by the ethics committee at Charité - Universitätsmedizin Berlin (no. EA2/157/22). The study was registered under DRKS00029907. Written informed consent was obtained from study participants before study start. The study was conducted in accordance with the declaration of Helsinki.

## 4 Results

### 4.1 Patient characteristics

There were 30 patients enrolled in the intervention group (21 of whom were women), and 15 patients enrolled in the control group (12 of whom were women). Detailed patient characteristics are presented in **Table 1**. Median disease duration was higher in the intervention group than in the control group (5.5 vs. 5.0 years). At baseline, clinical severity assessed by the current MGFA status was most commonly III (a or b) in both groups. MG-ADL scores were moderately higher in the intervention group compared to control group. The median QMG score in the intervention group was 15 points and 16 points in the control group.

**Table 1.** Patient characteristics at baseline.

| Characteristic | Overall<br>N=45 | Intervention<br>Group<br>N=30 | Control<br>Group<br>N=15 | Effect<br>Size | 95% CI |
| --- | --- | --- | --- | --- | --- |
| **Age, in years** |  |  |  | - | - |
| Median [IQR] | 48.0 [42.0, 58.0] | 47.5 [40.5, 57.5] | 48.0 [45.0, 56.5] |  |  |
| (Min, Max) | (23.0, 82.0) | (23.0, 82.0) | (25.0, 64.0) |  |  |
| **Disease duration, in years** |  |  |  | 0.2<sup>a</sup> | -0.18 to 0.5 |
| Median [IQR] | 5.0 [2.0, 9.0] | 5.5 [2.0, 9.8] | 5.0 [1.5, 8.0] |  |  |
| (Min, Max) | (1.0, 17.0) | (1.0, 17.0) | (1.0, 13.0) |  |  |
| **Sex** |  |  |  | - | - |
| Female, n (%) | 33 (73.3) | 21 (70.0) | 12 (80.0) |  |  |
| Male, n (%) | 12 (26.7) | 9 (30.0) | 3 (20.0) |  |  |
| **MGFA status at baseline, n (%)** |  |  |  | - | - |
| I | 1 (2.2) | 1 (3.3) | 0 (0.0) |  |  |
| IIa | 8 (17.8) | 5 (16.7) | 3 (20.0) |  |  |
| IIb | 9 (20.0) | 6 (20.0) | 3 (20.0) |  |  |
| IIIa | 6 (13.3) | 3 (10.0) | 3 (20.0) |  |  |
| IIIb | 15 (33.3) | 10 (33.3) | 5 (33.3) |  |  |
| IVa | - | - | - |  |  |
| IVb | 6 (13.3) | 5 (16.7) | 1 (6.7) |  |  |
| **MG-ADL score** |  |  |  | 0.2<sup>a</sup> | -0.1 to 0.5 |
| Median [IQR] | 9.0 [5.0, 11.0] | 10.0 [5.3, 11.0] | 7.0 [5.5, 9.5] |  |  |
| (Min, Max) | (1,14) | (1,14) | (4,14) |  |  |
| **QMG score** |  |  |  | -0.2<sup>a</sup> | -0.5 to 0.2 |
| Median [IQR] | 15.0 [13.0, 17.0] | 15.0 [12.3, 17.0] | 16.0 [14.5, 16.5] |  |  |
| (Min, Max) | (1, 25) | (1, 25) | (11, 25) |  |  |
| **Baseline therapies, n (%)**<br>(multiple answers possible) |  |  |  |  |  |
| **Pyridostigmine** | 42 (93.3) | 28 (93.3) | 14 (93.3) | 1.0<sup>b</sup> | 0.0 to 20.8 |
| **Glucocorticoids** | 26 (57.8) | 18 (60.0) | 8 (53.3) | 1.3<sup>b</sup> | 0.3 to 5.4 |
| **Immunosuppressive therapies\*** | 27 (60.0) | 17 (56.7) | 10 (66.7) | 0.7<sup>b</sup> | 0.1 to 2.8 |
| **Intensified treatment\*\*** | 27 (60.0) | 17 (56.7) | 10 (66.7) | 0.7<sup>b</sup> | 0.1 to 2.8 |
| **Intensified treatment in detail**<br>(multiple answers possible), n (%) |  |  |  |  |  |
| - **IVIg** | 14 (51.9) | 9 (52.9) | 5 (50.0) |  |  |
| - **PPH** | 6 (22.2) | 4 (23.5) | 2 (20.0) |  |  |
| - **Efgartigimod** | 4 (14.8) | 3 (17.6) | 1 (10.0) |  |  |
| - **Eculizumab** | 3 (11.1) | 2 (11.8) | 1 (10.0) |  |  |
| - **Ravulizumab**<sup>1</sup> | 6 (22.2) | 3 (17.6) | 3 (30.0) |  |  |
| - **Rituximab**<sup>2</sup> | 5 (18.5) | 5 (29.4) | 0 (0.0) |  |  |

\*azathioprine, mycophenolate mofetil, methotrexate (MTX)
\*\* in the last 12 months; multiple treatments at same time possible e.g. continuation of rituximab treatment and need of rescue therapy with PPH.220 <sup>a</sup> Cliff's Delta, <sup>b</sup> Odds Ratio, <sup>c</sup> Cramér's V221 <sup>1</sup> N=44; 1 missing in intervention group <sup>2</sup> N=44; 1 missing in control group

For all considered medical treatment options, there was no statistically significant difference between groups. At baseline, symptomatic therapy with pyridostigmine was most common (93.3%). More than half of the patients in each group were treated with either glucocorticoids (intervention group 60%; control group 53.3%), standard immunosuppressive therapy (intervention group 56.7%; 66.7% and/or received a form of intensified treatment in the past 12 months (intervention group 56.7%; control group 66.7%).

One intervention group patient withdrew from the study due to non-MG related reasons (experiencing a severe ocular infection resulting in significant visual impairment and prolonged hospitalization).

### 4.2 Telemedical communication patterns

#### Quantitative analysis of interaction days, contacts and topics

Mean study participation was 83.1 days. In the intervention group a total of 205 contacts were initiated by patients (**Table 2**). The median number of contacts per patient was six (IQR: 3, 8, range 1-22). Patients with the highest number of requests during the study (N=4), compared to the remaining intervention group (N=26), were older (67.0 vs. 47.0 years), and less clinically affected (MG-ADL score at baseline: 5 vs. 10.5 points). However, they more frequently addressed non-MG-related topics such as “Information regarding non-MG associated therapy” (26.9% vs. 16.2%) and “Non-MG related symptoms” (11.5% vs. 4.0%) (**Figure 1**). Interaction days ranged between 1 and 27 with a median of 9 interaction days (IQR: 5, 12). Specialists initiated contact with patients 274 times. Interaction days per patient by specialists ranged between 2 and 25 (**Table 2**).

**Figure 1.**
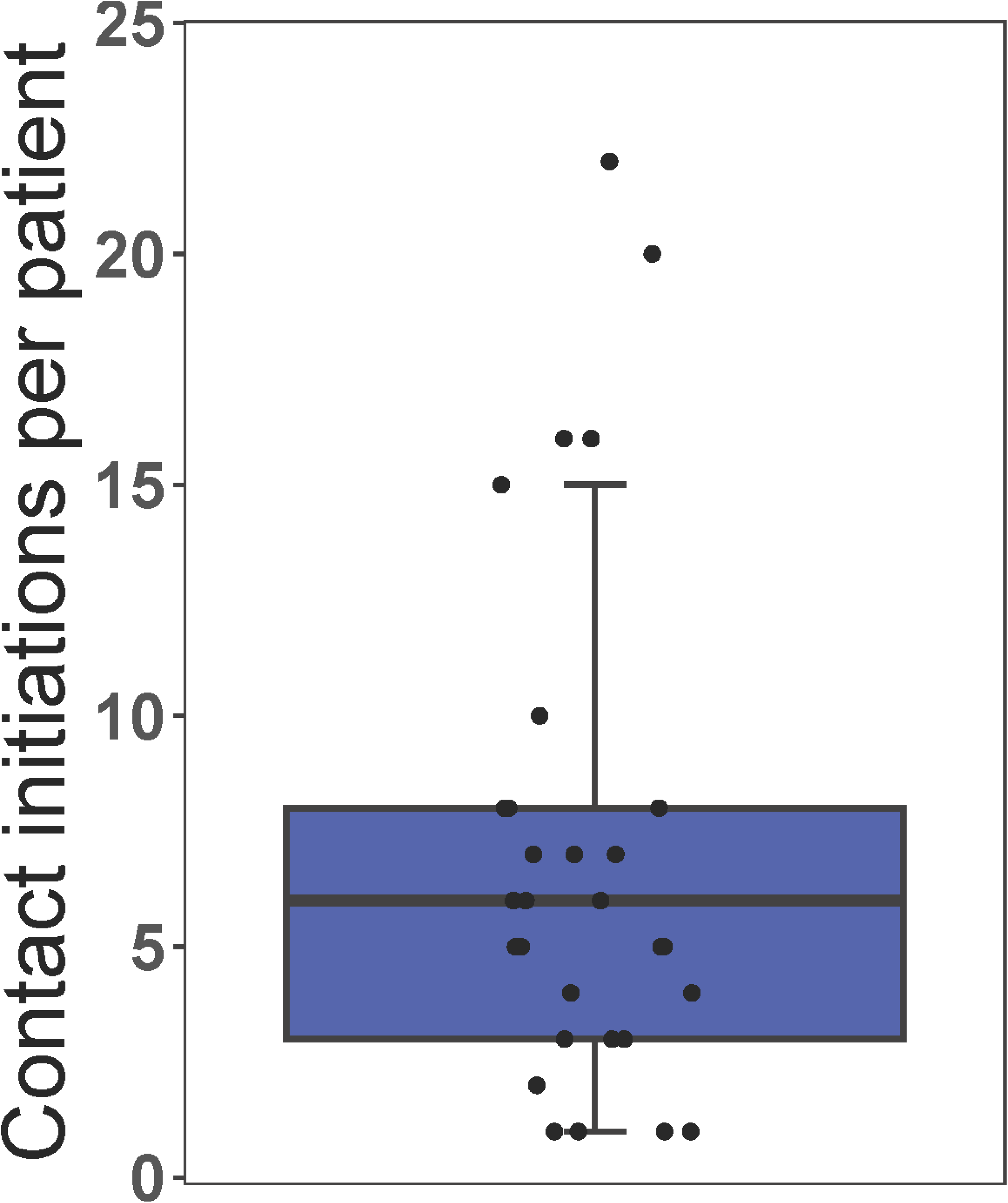
Number of contact initiations per patient during 12 weeks of study duration. (Median: 6, IQR: [3,8], (Min, Max): (1, 22))

**Table 2.** Quantitative analysis of contacts, topics, interactions.

|  | Patients | Specialists |
| --- | --- | --- |
| **<u>Contact initiations in total</u>** | **205<sup>1</sup>** | **274** |
| *Per patient* |  |  |
| Median [IQR] | 6.0 [3, 8] | 8.0 [6, 10.8] |
| (Min, Max) | (1, 22) | (1, 24) |
| **<u>Topics in total</u>** | **277** | **385** |
| *Per patient* |  |  |
| Median [IQR] | 7 [4, 12] | 10 [8, 17.5] |
| (Min, Max) | (1, 25) | (1, 38) |
| **<u>Interaction days in total</u>** | **278** | **307** |
| *Per patient* |  |  |
| Median [IQR] | 9 [5, 12] | 10 [7, 13] |
| (Min, Max) | (1, 27) | (2, 25) |

<sup>1</sup> with respect to N=29 proactive patients

#### Descriptive analysis of topics

##### Patient-initiated contacts

Overall, 277 patient contacts that contained distinct topics of interest were identified. The most frequently raised topics were *medical issues* (54.5%) followed by *technical inquiries* (25.3%), and *organizational inquiries* (19.1%) (**Table 3)**. The number of technical questions did not correlate with age (data not shown).

**Table 3.** Descriptive analysis of topics for patient-initiated contacts.

| Topics when patient contacts the specialist, N=277 | n (%) |
| --- | --- |
| **<u>Medical issues (overall)</u>** | **151 (54.5)** |
| MG-related therapy | 52 (34.4) |
| Myasthenic complaints | 37 (24.5) |
| Side effects of medication | 15 (9.9) |
| Information regarding non-MG associated therapy | 30 (19.9) |
| Non-MG related symptoms | 10 (6.6) |
| Other medical reasons | 7 (4.6) |
| **<u>Organizational inquiries</u>** | **53 (19.1)** |
| **<u>Technical inquiries (overall)</u>** | **70 (25.3)** |
| Initial setup | 6 (8.6) |
| Technical questions | 64 (91.4) |
| **<u>Other reasons<sup>1</sup></u>** | **3 (1.1)** |

<sup>1</sup>i.e. app-related feedback

#### Specialist-initiated contacts or recommendations

A total of 385 specialist contacts that contained distinct topics were identified, 337 in between TCUs and 48 at TCUs (**Table 4**). Technical issues presented especially at the start of the study (**Supplemental Figure 1**). The topics addressed by specialists in between TCUs were: *medical issues* (45.1%), *technical issues* (44.2%), and *organizational issues* (10.7%)*. Change in medication* (41.1%) was the most frequent subtopic of *medical issues*. At TCUs, the most frequent topic addressed by specialists was *medical issues* (79.2%)*, most frequently deterioration of monitoring data* (31.6%), followed by *consultation about clinical complaints* (28.9%), and *change in medication* (26.3%). Specialists contacted patients in 49.2% of TCUs after performing a systematic data evaluation. In more than half of contacts PROMs (54.5%) were relevant trigger factors for contact initiation (multiple trigger factors per contact possible) (**Tables 4 and 5**). 25% of physician-initiated contacts were prompted by deterioration in monitoring data, an even higher proportion (75%) were based solely on information from PROMs and wearable data, without any preceding patient-reported complaints in the chat.

**Table 4.**
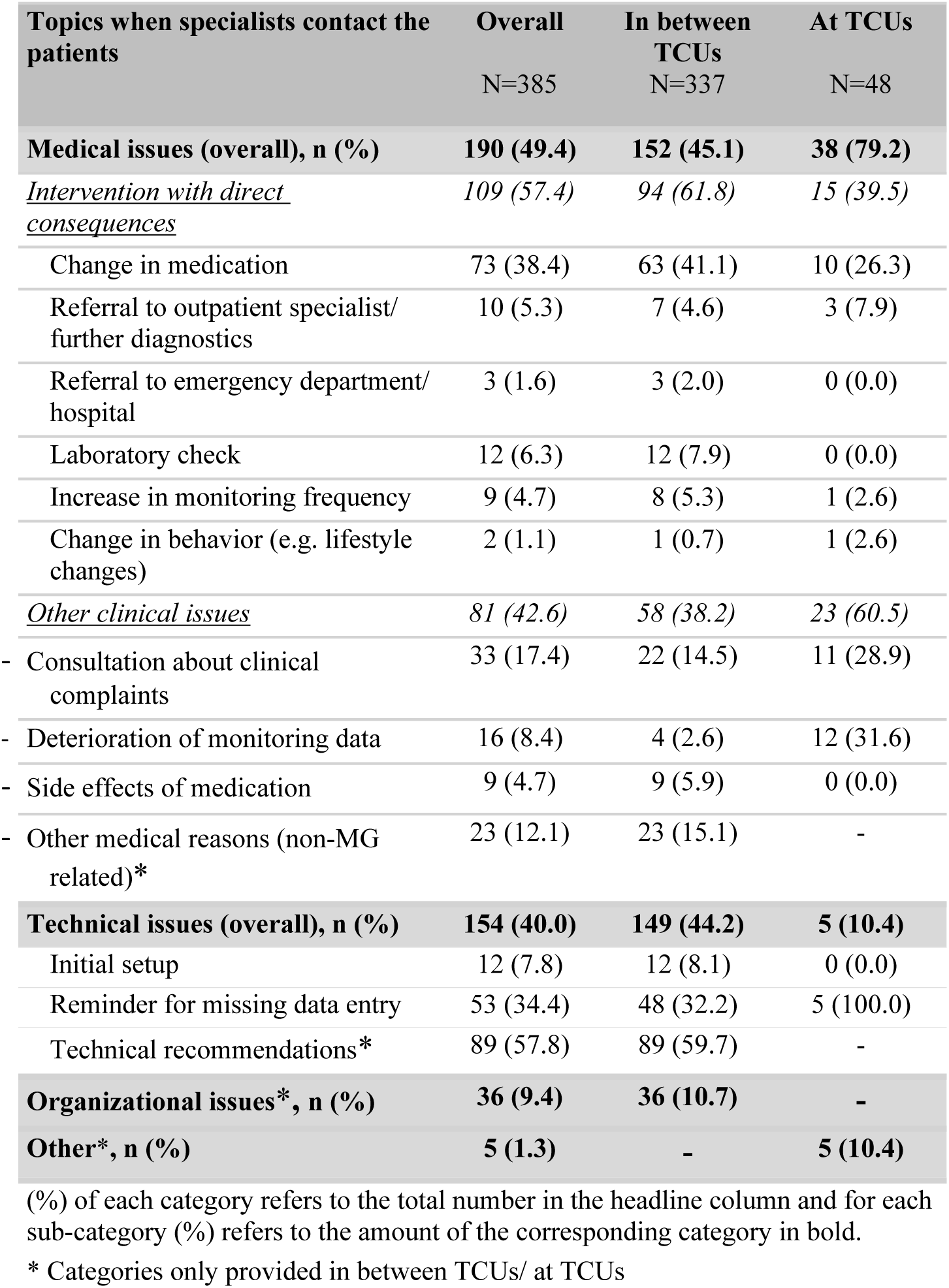
Descriptive analysis of topics addressed by specialists.

| Topics when specialists contact the patients | Overall<br>N=385 | In between TCUs<br>N=337 | At TCUs<br>N=48 |
| --- | --- | --- | --- |
| **Medical issues (overall), n (%)** | **190 (49.4)** | **152 (45.1)** | **38 (79.2)** |
| *Intervention with direct consequences* | *109 (57.4)* | *94 (61.8)* | *15 (39.5)* |
| Change in medication | 73 (38.4) | 63 (41.1) | 10 (26.3) |
| Referral to outpatient specialist/<br>further diagnostics | 10 (5.3) | 7 (4.6) | 3 (7.9) |
| Referral to emergency department/<br>hospital | 3 (1.6) | 3 (2.0) | 0 (0.0) |
| Laboratory check | 12 (6.3) | 12 (7.9) | 0 (0.0) |
| Increase in monitoring frequency | 9 (4.7) | 8 (5.3) | 1 (2.6) |
| Change in behavior (e.g. lifestyle<br>changes) | 2 (1.1) | 1 (0.7) | 1 (2.6) |
| *Other clinical issues* | *81 (42.6)* | *58 (38.2)* | *23 (60.5)* |
| - Consultation about clinical<br>complaints | 33 (17.4) | 22 (14.5) | 11 (28.9) |
| - Deterioration of monitoring data | 16 (8.4) | 4 (2.6) | 12 (31.6) |
| - Side effects of medication | 9 (4.7) | 9 (5.9) | 0 (0.0) |
| - Other medical reasons (non-MG<br>related)* | 23 (12.1) | 23 (15.1) | - |
| **Technical issues (overall), n (%)** | **154 (40.0)** | **149 (44.2)** | **5 (10.4)** |
| Initial setup | 12 (7.8) | 12 (8.1) | 0 (0.0) |
| Reminder for missing data entry | 53 (34.4) | 48 (32.2) | 5 (100.0) |
| Technical recommendations* | 89 (57.8) | 89 (59.7) | - |
| **Organizational issues\*, n (%)** | **36 (9.4)** | **36 (10.7)** | **-** |
| **Other\*, n (%)** | **5 (1.3)** | **-** | **5 (10.4)** |

(%) of each category refers to the total number in the headline column and for each sub-category (%) refers to the amount of the corresponding category in bold.
\* Categories only provided in between TCUs/ at TCUs

**Table 5.** Reasons for specialists initiating contact with patients after telemedical check-ups.

|  | n (%) |
| --- | --- |
| Contact initiation by the specialist based on TCU (total TCUs N=59) | 29 (49.2) |
| **Reasons for specialists initiating contact with patients based on TCUs (multiple answers possible), Total Triggers N=99** | **n (%)** |
| *Information based on chat interactions* | 11 (11.1) |
| *Medical reports and documents (uploaded in the app)* | 0 (0.0) |
| *PROMs* | 54 (54.5) |
| MG-ADL | 12 (12.1) |
| MG-QoL15r | 9 (9.1) |
| SSQ | 12 (12.1) |
| PASS | 3 (3.0) |
| HADS | 8 (8.1) |
| Chalder Fatigue Scale | 10 (10.1) |
| *Wearables* | 17 (17.2) |
| Heart rate | 2 (2.0) |
| Oxygen saturation | 7 (7.1) |
| Step count | 8 (8.1) |
| *Spirometry (FVC)* | 8 (8.1) |
| *Single Breath Count Test (SBCT)* | 9 (9.1) |

### 4.3 Contact with other physicians concerning MG-related issues

Patients in the control group contacted other physicians more frequently than those in the intervention group but the difference was not significant (**Table 6**). In both groups, outpatient neurologists were most frequently contacted.

**Table 6.**
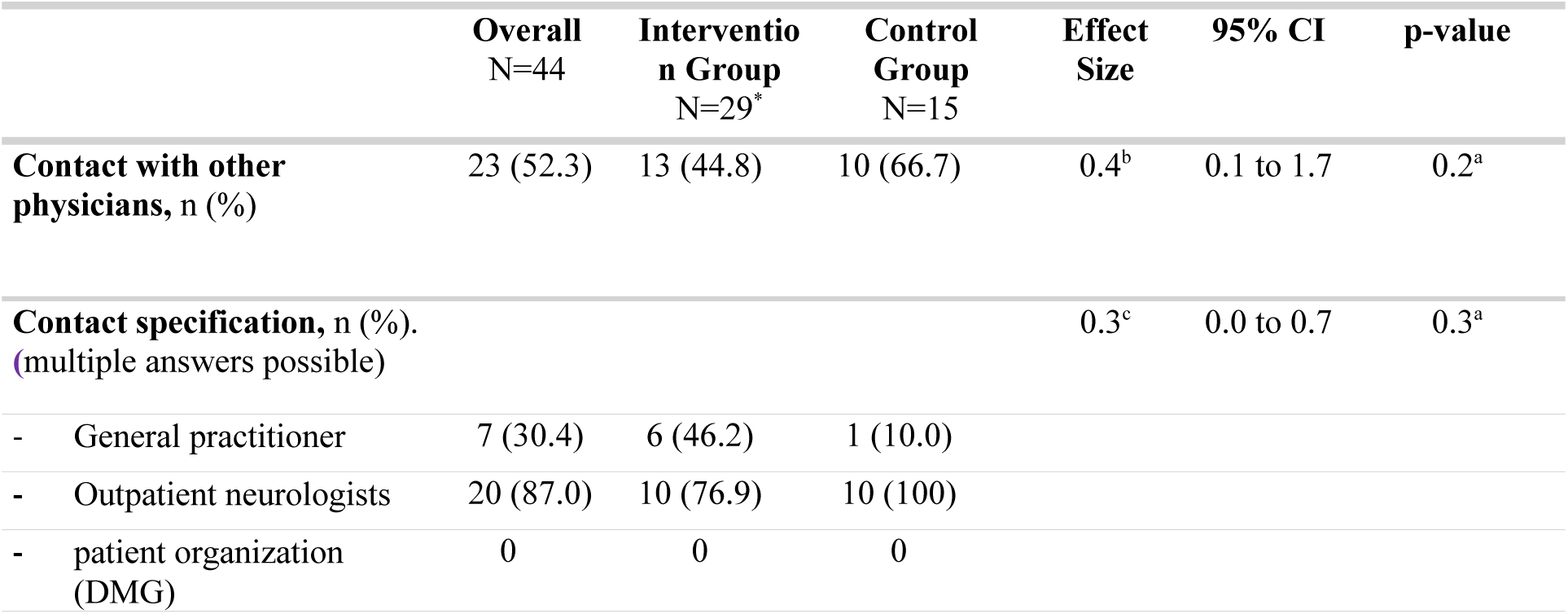
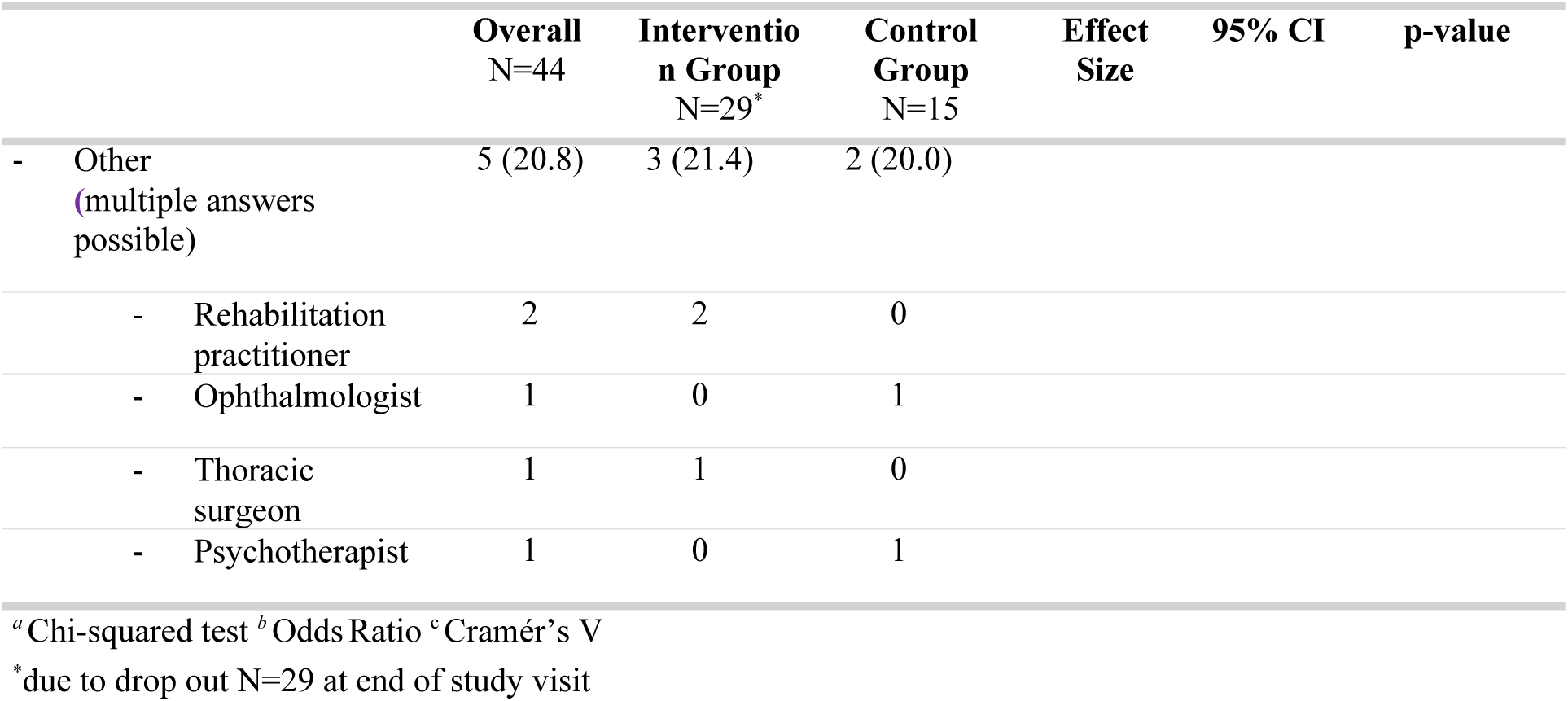
Contact with other physicians for MG-related issues throughout the study.

### 4.4 Satisfaction with specialized treatment availability

At the baseline visit there was no statistically significant difference between intervention group and control group in proportions of responses to the question “I have the feeling that I can reach a specialist in an adequate amount of time if my MG symptoms worsen”. At the end of the study all patients in the intervention group (100%) agreed (“yes”) with the following prompt “I have the feeling that with MyaLink I can reach a specialist in an adequate amount of time if my MG symptoms worsen.”. This was significantly different from the baseline response, thus indicating a strong association between timepoints (before and after using MyaLink) and response (Eta-squared η^2^=0.5; p<0.1) (**Figure 2; Supplemental Table 1**).

**Figure 2.**
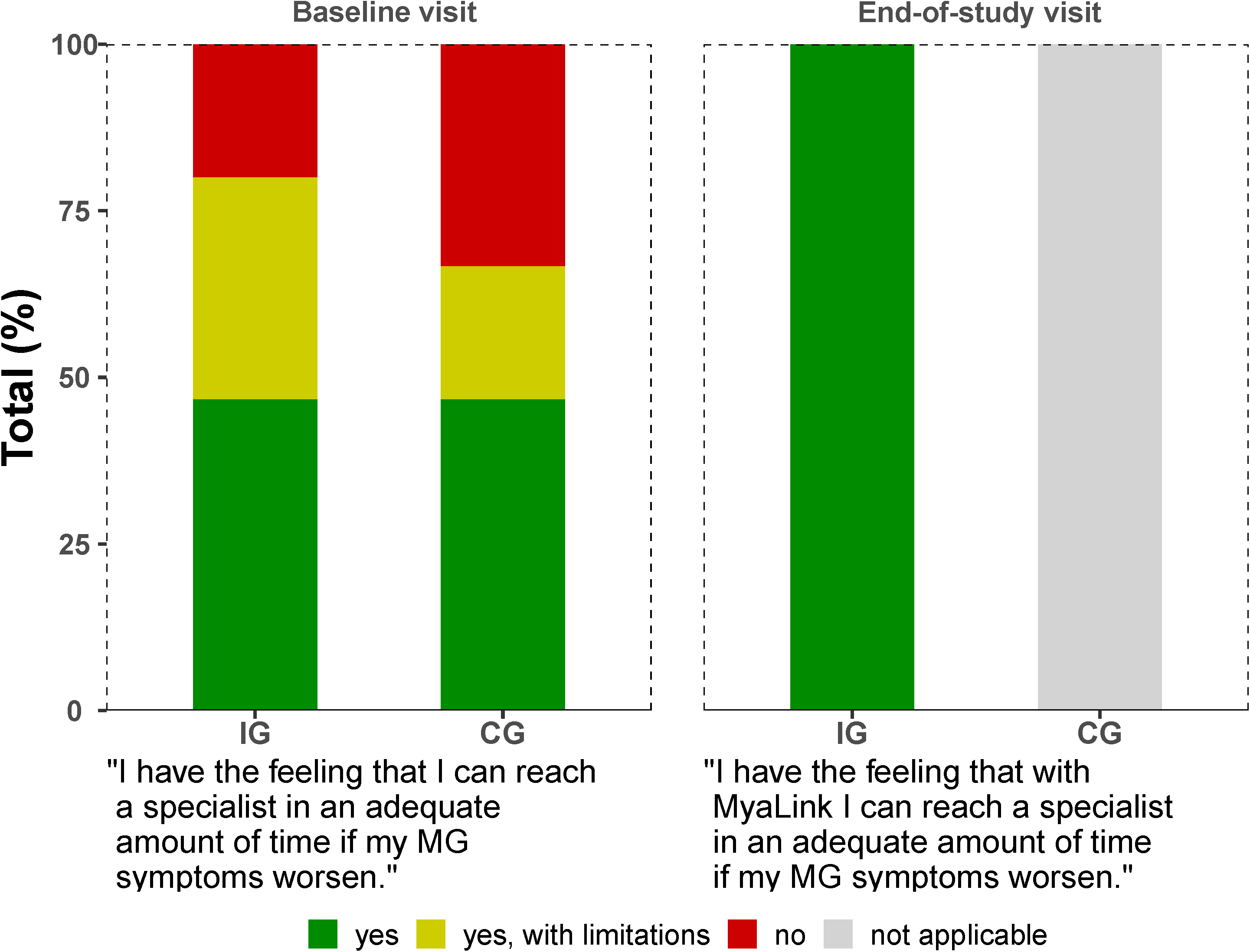
Perceived **accessibility of MG specialists** in the control and intervention group at baseline visit (left) and at the end-of study visit (right)

## 5 Discussion

In this study, telemedical communication patterns between MG patients and specialists were assessed.

More than half of the topics addressed both by the patients and specialists pertained to medical issues. Interestingly, 57.4% of specialist recommendations on medical topics required direct interventions e.g., changes in medication. We hypothesize that this indicates a medical need that is unlikely to be met in a timely manner without telemedical intervention. Comparison with a study analyzing MG patient inquiries via email showed a similar level of medical inquiries.^6^ Notably, there was a higher number of questions concerning organizational issues addressed to the center through e-mail (30%) compared to 19.1% in this study. One possible reason could be the high number of inquiries for appointments and medical records that arose in the e-mail analysis which were not common in our study as follow-up appointments had been planned preemptively.

In the intervention group the median of interaction days was greater than the number of patient-specialist interactions typically observed in standard care at the iMZ with only in-person appointments. Limited expert opinion could lead to patients seeking information about MG from patient organizations or through online health communities.^27^ However, online medical misinformation has been critically discussed previously.^28–31^

When performing structured TCUs, the initiation of contact was majorly triggered solely by a combination of remote monitoring parameters indicating an increase in symptoms, most commonly including PROMs. These observations emphasize the added value of telemonitoring in uncovering relevant clinical changes that may not yet be perceived or reported by patients. Although MG-specific PROMs are established outcome parameters in clinical studies,^32^ there is limited data on the consistency of PROM evaluation and how it influences decision-making in clinical practice, despite expert consensus recommending their use. ^33^ One reason may be hesitations due to additional (time-consuming) work for healthcare personnel. However, a strong correlation between patient-assessed and physician-assessed MG-ADL has been shown thus limiting additional work for physicians.^34, 35^

In addition, data from wearables (activity tracker), spirometry and SBCT also prompted TCU contacts. Changes of these measurements may be predictive of impeding myasthenic exacerbation or crisis^36, 37^ and continuous monitoring of oxygen saturation may be valuable for identifying patients with sleep related breathing problems in MG.^38, 39^ Given that pronounced respiratory dysfunction is relatively rare in MG in general and in our cohort in particular, it is not surprising that such data did not trigger contacts more frequently in our study.

Particularly in rare diseases, accessible specialist care is an important factor for treatment satisfaction.^40^ In this study, patients felt accessibility of specialists was improved significantly through telemedical approach. In accordance with our results, MG patients were found to be generally open to telemedical consultations. ^41^ However, so far most mobile applications for rare diseases focus on self-monitoring or provide informative content.^42^ In MG in particular there have been studies on diagnosis of MG with telemedical tools^43, 44^, platforms combining symptom monitoring and expert support have mainly been reported outside of rare (neuromuscular) diseases.^45–48^

Although our telemedical approach improved access to specialist neurologist services, patients continued to visit other physicians for MG-related care, with no statistically significant differences observed between both groups. This may be due to the small sample size, as indicated by the wide confidence intervals, or the relatively short three-month study duration. Additionally, source data suggest that patients frequently consulted other physicians for routine follow-ups or pre-scheduled laboratory tests.

Furthermore, specialists had more interaction days and initiated more contacts than patients. In view of generally scarce resources on the practitioner side, such an effort in the communication tool seems high, especially because this is hardly ever remunerated. It remains unclear whether the time spent on remote communication and problem-solving can reduce the need for other resources, such as in-person visits or hospital admissions and should be investigated in future studies. Dividing interaction patterns into TCUs and in between TCUs we aimed to systematically differentiate between reactions to active patient-established contacts and specialist-established contacts that followed systematic review of monitoring data. There was a far smaller number of contacts at TCUs, indicating that through constant availability of medical advice larger issues may have been resolved throughout.

There are several limitations for this study. There was a relatively high number of contacts addressing technical problems. As this is a pilot study with a MG-tailored platform version technical problem were somewhat expected and they fortunately became less frequent over the course of the study. Future technical refinements are necessary and patient training will be important.

Further limitations include the relatively small cohort size and short study period. Patients with higher digital literacy may have been more likely to participate, potentially affecting the representativeness of the cohort . The regular monitoring and continuous collection of clinical data may have contributed to increased adherence and more frequent communication. In the intervention group there were more patients with a high MGFA status (IVb) at baseline and there was a relatively high percentage of patients overall that had received intensified treatment in the past twelve months indicating a relatively severely affected study group. In the control group, informal communication with the iMZ that may have occurred (e.g. via publicly available phone numbers or email addresses) but was not part of standard of care was not captured systematically thus limiting insight into this patient group. Patients from the control group were not questioned about accessibility of specialized MG treatment at the end-of-study visit which may have potentially been influenced by study participation even in the control group.

## Conclusions

Our results reveal a relevant need for timely advice especially concerning treatment recommendations in patients with MG. PROMs and data from external devices aided in medical decision making and patient-specialist interaction through a telemedical solution using MyaLink was valued and accepted. Patients reported a significant improvement in perceived accessibility of specialist care. Future studies are needed to evaluate the potential of telemedical solutions for MG, particularly for exacerbation and crisis prevention and their implementation in clinical care processes.

## Acknowledgements

We thank the NeuroScience Clinical Research Center at Charité – Universitätsmedizin Berlin for their support in administration and throughout the formal processes essential for the successful execution of this study. The authors especially wish to thank Maja Olzewska, Josefina Kreusler, and Gabriela Jiron-Lozano for their support in establishing the REDCap® database. Furthermore, special acknowledgements are extended to Jens Brestrich and Dike Remstedt for their support in conducting the study. Additionally, the authors thank Qurasoft GmbH for providing the software for the study, namely Lale Zils from Qurasoft GmbH for the project assistance during the study and Sebastian Thunert, Lucas Keune and Ragnar Hirschbrich from the Qurasoft team for their technical support throughout the study. We are grateful for the participation of patients and for the many constructive discussions with DMG e.V. during the platform’s development and all study phases. Their feedback and suggestions were very valuable in improving the platform and ensuring a patient-centered perspective. We wish to thank all SP that participated in the study.

## Author contributions

**M.H. and S.L.:** Writing- Original draft preparation. **M.S., S.L. and L.G.:** Conceptualization, Methodology. **A.M.:** Supervision, Resources. **M.S., S.L., D.L.**: Investigation. **M.S., S.L. and L.G.:** Validation. **M.M:** Formal analysis, Visualization. **M.H., S.L., M.M., L.G., D.L., P.N., H.S., A.M., M.S.:** Writing- Reviewing and Editing.

## Statements and Declarations

### Ethical considerations

This study received approval by the ethics committee at Charité - Universitätsmedizin Berlin (no. EA2/157/22). The study was registered under DRKS00029907. The study was conducted in accordance with the declaration of Helsinki.

### Consent to participate

Written informed consent was obtained from study participants before study start.

### Consent for publication

Written informed consent was obtained from study participants before study start.

### Data availability

Data not provided in the article may be shared (anonymized) upon request from the corresponding author for purposes of replicating procedures and results.

## Abbreviations

DMG: German Myasthenia Association
ICD-10: International statistical classification of diseases and related health problems 10th revision
MG: Myasthenia gravis
MGFA: Myasthenia gravis foundation of America
MG-ADL: Myasthenia gravis activities of daily living
SBCT: Single breath count test
QMG: Quantitative Myasthenia gravis
IVIg: Intravenous immunoglobulins
PPH: Plasmapheresis
PROMS: Patient-Reported Outcome Measures
TCU: Telemedical check-up
FVC: Forced vital capacity

