## supplemental for "Telemedical communication patterns in myasthenia gravis in a remote monitoring study"

**Supplemental material**

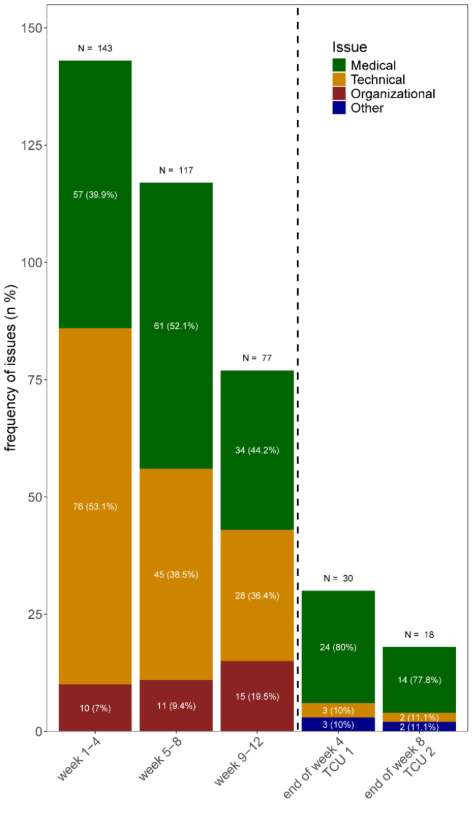

***Supplement Figure 1*** *Topics addressed by specialists through the chat at different time points during study period.*

|  | **Overall**  N=45 | | **Intervention group**  N=30 | **Control  group**  N=15 | **p-value\| Effect Size 95%-CI** |
| --- | --- | --- | --- | --- | --- |
| **Assessed during baseline visit, n (%)**  *“I have the feeling that I can reach a specialist in an adequate amount of time if my MG symptoms worsen.”* | | | | | 0.5^a^ \| 0.2^b^ (0.0 to 0.5) |
| Yes | 21 (46.7) | | 14 (46.7) | 7 (46.7) |  |
| Yes with limitations | 13 (28.9) | | 10 (33.3) | 3 (20.0) |  |
| No | 11 (24.4) | | 6 (20.0) | 5 (33.3) |  |
| **Assessed during end-of-study visit, 1 missing, n (%)**  *“I have the feeling that with MyaLink I can reach a specialist in an adequate amount of time if my MG symptoms worsen.”* | | | | |  |
| Yes | |  | 29 (100.0) |  |  |
| Yes with limitations | |  | 0 (0.0) |  |  |
| No | |  | 0 (0.0) |  |  |
| **p-value\| Effect Size** | |  | <0.1^c^ \| 0.5^d^ |  |  |
| ^a^ Pearson’s Chi-Squared test ^b^ Cramér’s V | | | | | |
| ^c^ Cochran’s Q test ^d^ Eta-squared (η^2^) | | | | | |

***Supplement Table 1*** *Availability of specialized medical services for the treatment of Myasthenia gravis.*
